# Utility of Genome Sequencing After Nondiagnostic Exome Sequencing in Unexplained Pediatric Epilepsy

**DOI:** 10.1101/2024.08.08.24307445

**Authors:** Alissa M. D’Gama, Wanqing Shao, Lacey Smith, Hyun Yong Koh, Maya Davis, Julia Koh, Brandon T. Oby, Cesar I. Urzua, Beth R. Sheidley, Shira Rockowitz, Annapurna Poduri

## Abstract

**Importance:** Epilepsy is the most common neurological disorder of childhood. Identifying genetic diagnoses underlying epilepsy is critical to developing effective therapies and improving outcomes. Most children with non-acquired (unexplained) epilepsy remain genetically unsolved, and the utility of genome sequencing after nondiagnostic exome sequencing is unknown.

**Objective:** To determine the diagnostic (primary) and clinical (secondary) utility of genome sequencing after nondiagnostic exome sequencing in individuals with unexplained pediatric epilepsy.

**Design:** This cohort study performed genome sequencing and comprehensive analyses for 125 participants and available biological parents enrolled from August 2018 to May 2023, with data analysis through April 2024 and clinical return of diagnostic and likely diagnostic genetic findings. Clinical utility was evaluated.

**Setting:** Pediatric referral center

**Participants:** Participants with unexplained pediatric epilepsy and previous nondiagnostic exome sequencing; biological parents when available

**Exposure(s):** Short-read genome sequencing and analysis

**Main Outcome(s) and Measure(s):** Primary outcome measures were the diagnostic yield of genome sequencing, defined as the percentage of participants receiving a diagnostic or likely diagnostic genetic finding, and the unique diagnostic yield of genome sequencing, defined as the percentage of participants receiving a diagnostic or likely diagnostic genetic finding that required genome sequencing. The secondary outcome measure was clinical utility of genome sequencing, defined as impact on evaluation, treatment, or prognosis for the participant or their family.

**Results:** 125 participants (58 [46%] female) were enrolled with median age at seizure onset 3 [IQR 1.25, 8] years, including 44 (35%) with developmental and epileptic encephalopathies. The diagnostic yield of genome sequencing was 7.2% (9/125), with diagnostic genetic findings in five cases and likely diagnostic genetic findings in four cases. Among the solved cases, 7/9 (78%) required genome sequencing for variant detection (small copy number variant, three noncoding variants, and three difficult to sequence small coding variants), for a unique diagnostic yield of genome sequencing of 5.6% (7/125). Clinical utility was documented for 4/9 solved cases (44%).

**Conclusions and Relevance:** These findings suggest that genome sequencing can have diagnostic and clinical utility after nondiagnostic exome sequencing and should be considered for patients with unexplained pediatric epilepsy.

**Key Points:** *Question:* What is the utility of genome sequencing after nondiagnostic exome sequencing in individuals with unexplained pediatric epilepsy?

*Findings:* In this cohort study of 125 individuals with unexplained pediatric epilepsy and nondiagnostic exome sequencing, genome sequencing identified diagnostic genetic findings in five cases and likely diagnostic genetic findings in four cases. Of the nine solved cases, seven required genome sequencing to solve, and four had documented clinical utility.

*Meaning:* Genome sequencing can identify genetic diagnoses not detectable by exome sequencing and should be considered for participants with unexplained pediatric epilepsy, as first-line testing or after nondiagnostic exome sequencing.

## Introduction

Epilepsy, defined as recurrent unprovoked seizures, has a 1 in 26 lifetime prevalence and is the most common pediatric neurological disorder worldwide.^1–3^ Pediatric epilepsies are associated with substantial morbidity, notably drug-resistant seizures in one in three patients, developmental delay (DD), intellectual disability (ID), and autism spectrum disorder (ASD).^2,4–6^ Identifying the etiologies underlying pediatric epilepsies is critical to developing etiology-based therapies and ultimately improving outcomes.

Many pediatric epilepsies are non-acquired (“unexplained”) and have presumed molecular genetic etiologies.^7–11^ Identifying an underlying genetic etiology ends the diagnostic odyssey and may inform clinical management, including workup and treatment, as well as prognosis, genetic counseling, and eligibility for etiology-based research, notably natural history studies and clinical trials of emerging precision therapies.^12^ However, most unexplained pediatric epilepsies remain genetically unsolved.^13–16^ A recent systematic evidence review (SER) reported the diagnostic yield of chromosomal microarray (CMA) as 9%, multi-gene panels 19%, and exome sequencing (ES) 24%.^14^ While CMA and panels were early first-line genetic tests in clinical epilepsy care, ES has increasingly been used, including as first-line, when covered by payors.

Given >800 reported monogenic etiologies of epilepsy, comprehensive genetic testing is necessary to maximize diagnostic yield.^17^ Short-read genome sequencing (GS) has the potential to identify nearly all pathogenic variants detectable by other approaches as well as novel pathogenic variants uniquely detectable by GS.^18^ Compared to ES, which targets coding regions (∼2% of the genome), GS provides more uniform coverage and has the potential to detect single nucleotide variants (SNVs) and small insertions-deletions (indels) in coding regions that are missed by ES. GS also has the potential to detect additional copy number variants (CNVs) and structural variants (SVs), short tandem repeats (STRs), mobile element insertions (MEIs), mitochondrial variants, and noncoding variants that are difficult or not possible to detect by ES.

There have been limited studies of GS in epilepsy cohorts; the SER identified only four studies with overall 48% diagnostic yield.^14^ As GS is more expensive than ES and requires more time and effort to analyze, it is important to understand the additional utility of GS compared to ES to inform optimal implementation of GS in clinical epilepsy care. Recent evidence-based guidelines from the National Society of Genetic Counselors and endorsed by the American Epilepsy Society recommend genetic testing for all patients with unexplained epilepsy, with ES/GS conditionally recommended over panels for first-tier testing.^19^ Two small studies (n=15 and n=20) in highly selected cohorts (developmental and epileptic encephalopathies [DEEs] and epileptic encephalopathies [EEs]) have reported the diagnostic yield of GS after nondiagnostic ES, and did not perform analyses for all variant types detectable by GS.^20,21^ Thus, the true benefit of GS compared to ES in unexplained pediatric epilepsy remains unknown. Here, we evaluate the utility of GS after nondiagnostic ES in a larger (n=125, including n=44 with DEE/EE) and rigorously phenotyped unexplained pediatric epilepsy cohort using comprehensive analyses.

## Methods

### Ethics

This cohort study was approved by the Boston Children’s Hospital (BCH) Institutional Review Board. Participants or their guardians provided written informed consent. Data were reported according to the Strengthening the Reporting of Observational Studies in Epidemiology guideline.

### Study Cohort

Between August 2018 and May 2023, we recruited patients with unexplained epilepsy from BCH, and biological parents and affected siblings whenever possible. We excluded patients with known acquired or genetic cause for epilepsy. For participants without previous clinical ES, we first performed research ES and analyses for SNVs, indels, and CNVs, with a diagnostic yield of 19.2% as previously reported.^13^ For participants who remained genetically unsolved after clinical and/or research ES, we performed research GS.

### Phenotyping

Clinical data were abstracted from the electronic medical record (EMR) and stored in a BCH-hosted Research Electronic Data Capture database.^22^ Data collected included sex, parent-reported race and ethnicity, age at seizure onset, EEG findings, MRI findings, other clinical features, previous genetic testing, and family medical history. We categorized age of seizure onset as previously described.^13^ Epilepsy syndromes were classified according to International League Against Epilepsy definitions,^5,6,23–25^ with each participant categorized as DEE, EE, genetic generalized epilepsy (GGE), non-acquired focal epilepsy (NAFE), or combined generalized and focal epilepsy (Combined).

### GS and Analyses

GS and analyses were completed through the Children’s Rare Disease Collaborative.^26^ Blood samples from participants were collected and shipped to a Clinical Laboratory Improvement Amendments (CLIA) certified laboratory (GeneDx, Gaithersburg, MD) for DNA extraction and GS. Sequencing data were securely transferred to BCH and analyses were performed using various computational workflows (further details in eMethods in the Supplement).^26–32^

### Variant Classification

Candidate variants were reviewed by a multidisciplinary epilepsy genetics team and classified based on American College of Medical Genetics and Genomics, Association for Molecular Pathology, and Clinical Genome Resource guidelines.^33,34^ The diagnostic yield includes 1) participants with diagnostic genetic findings, defined as Pathogenic (P)/Likely Pathogenic (LP) variants impacting genes associated with human disease that fit the participant phenotype and disease mode of inheritance and 2) participants with likely diagnostic genetic findings, defined as Variants of Uncertain Significance (VUS) impacting genes associated with human disease that fit the participant phenotype and disease mode of inheritance and deemed likely diagnostic after further review of clinical data and/or investigations.

### Unique to GS Framework

We considered the following variant types unique to GS, i.e., requiring GS and unable to be detected using current ES methods: 1) noncoding variants, 2) coding SNVs and indels not called on ES analyses, 3) CNVs smaller than three exons, 4) other SVs not called on ES analyses, 5) STRs not called on ES analyses, and 6) mitochondrial genome variants.

### Return of Results

For participants who consented to receive results, clinical confirmation of diagnostic/likely diagnostic genetic findings was performed using DNA maintained at the CLIA-certified laboratory (GeneDx) when the laboratory had a clinically accredited test available. Clinical confirmation reports were uploaded to the EMR and results returned to families by a physician and genetic counselor through the BCH Epilepsy Genetics Program.

### Clinical Utility

Clinical utility after return of results was abstracted from the EMR and defined as previously described: influence on treatment, potential for precision therapy, indication or avoidance of additional investigations, additional prognostic information, influence on goals of care, or influence on genetic counseling beyond recurrence risk, which was discussed with all families during return of results.^35^

### Statistical Analysis

Summary statistics were analyzed for cohort demographics and clinical features. The diagnostic yield of GS was calculated as the percentage of participants who received a diagnostic or likely diagnostic genetic finding. The unique diagnostic yield of GS was calculated as the percentage of participants who received a diagnostic or likely diagnostic genetic finding that required GS. The clinical utility of GS was calculated as the percentage of participants with a diagnostic or likely diagnostic genetic finding for which the diagnosis had clinical utility.

## Results

### Cohort Demographics

Our cohort included 125 individuals with pediatric-onset epilepsy and nondiagnostic ES; 58 (46%) were female (**Table 1**). Most (113/125 [90%]) had ES data available in the research setting; only 12/125 (10%) had ES performed clinically without data available in the research setting. Per parent report, 96 individuals (77%) were White, 7 (6%) Asian, 4 (3%) Black, 3 (2%) Middle Eastern/North African, and 15 (12%) had unknown race; 111 (89%) were non-Hispanic, 10 (8%) Hispanic, and 4 (3%) had unknown ethnicity. Median age at seizure onset was 3 years (IQR 1.25-8), with 27 (22%) with onset in the first year. Forty-four participants (35%) had DEE/EE (43 DEE, one EE). Of the participants without DEE/EE, 30 (24% of the total cohort) had NAFE, 33 (27%) GGE, and 18 (14%) combined focal and generalized epilepsy. We had trio GS data for 49 participants (39%), duo GS data for 19 (15%, all with ES data from at least one biological parent available), and proband GS data for 57 (46%, 52 (91%) with ES data from at least one biological parent available).

**Table 1:** Cohort Demographics.

|  | **Number (%)**<br>**(N=125)** |
| --- | --- |
| **Sex** | 67 (54) |
| Male | 58 (46) |
| Female |  |
| **Race** |  |
| White | 96 (77) |
| Asian | 7 (6) |
| Black | 4 (3) |
| Middle Eastern/North African | 3 (2) |
| Unknown | 15 (12) |
| **Ethnicity** |  |
| Non-Hispanic | 111 (89) |
| Hispanic | 10 (8) |
| Unknown | 4 (3) |
| **Age at seizure onset** |  |
| Median (IQR; years) | 3 (1.25,8) |
| Neonatal (<1 month) | 4 (3) |
| Infantile (1 to <12 months) | 23 (18) |
| Early Childhood (1 to <6 years) | 57 (46) |
| School-aged (6 to <14 years) | 34 (27) |
| Adolescent (>=14 years) | 7 (6) |
| **Epilepsy type** |  |
| *DEE<sup>a</sup>* | 44 (35) |
| *Non-DEE<sup>b</sup>* | 81 (65) |
| NAFE | 30 (24) |
| GGE | 33 (27) |
| Combined | 18 (14) |
| **Type of GS** |  |
| *Trio GS* | 49 (39) |
| *Duo GS* | 19 (15) |
| Trio ES | 8 (42) |
| Duo ES | 11 (58) |
| *Singleton GS* | 57 (46) |
| Trio ES | 38 (67) |
| Duo ES | 14 (24) |
| Singleton ES | 5 (9) |

<sup>a</sup>One participant with EE
<sup>b</sup>Thirty-seven participants with developmental delay, intellectual disability, and/or autism spectrum disorder

### Diagnostic Utility of GS

Short-read GS and comprehensive analyses were performed to identify coding SNVs and indels, CNVs, SVs, STRs, MEIs, mitochondrial, and noncoding variants (**Figure 1**). We identified diagnostic genetic findings in five and likely diagnostic genetic findings in four individuals, for a diagnostic yield of GS of 7.2% (9/125) (**Table 2**). This included 4/44 (9.1%) individuals with DEE and 5/81 (6.2%) with non-DEE. Of the nine solved cases, seven (78%) required GS for variant detection and could not be solved by current ES methods, for a unique diagnostic yield of GS of 5.6% (7/125) (**Figure 2**). This included 4/4 (100%) of the solved cases with DEE, for a unique yield of GS of 9.1% (4/44), and 3/5 (60%) of the solved cases with non-DEE, for a unique yield of GS of 3.7% (3/81).

**Figure 1:**
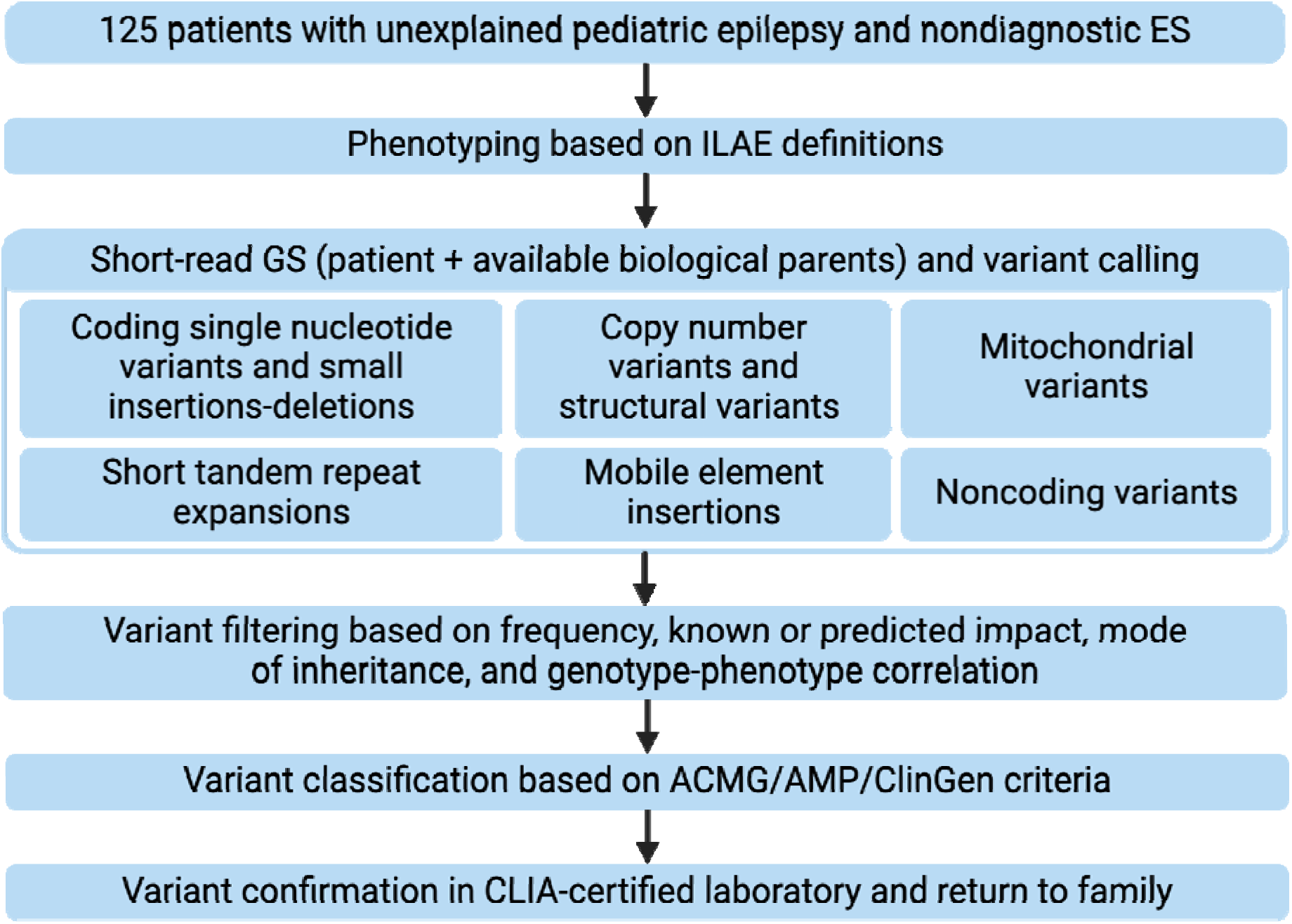
Genome Sequencing and Analysis Framework. We performed genome sequencing (GS) and comprehensive analysis for 125 patients with unexplained pediatric epilepsy and nondiagnostic ES. Epilepsy-focused phenotyping was performed using International League Against Epilepsy (ILAE) definitions and variant classification was performed using American College of Medical Genetics and Genomics, Association for Molecular Pathology, and Clinical Genome Resource (ACMG/AMP/ClinGen) criteria.

**Figure 2:**
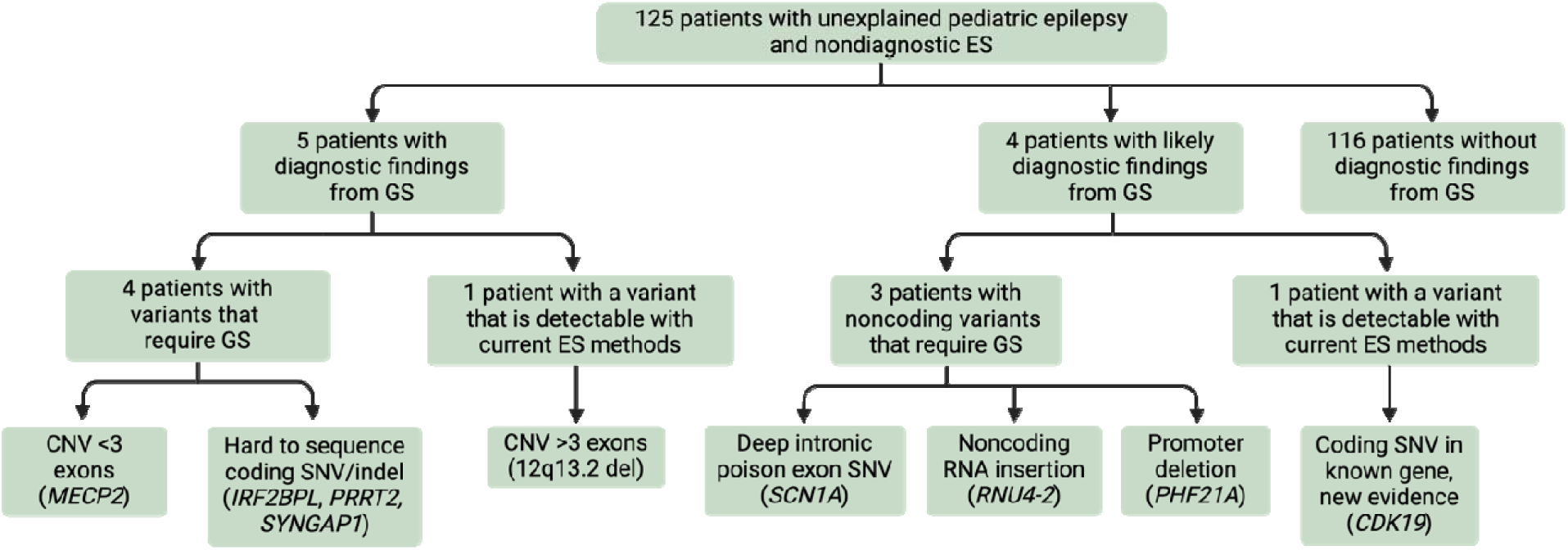
Study Workflow. In our cohort of 125 patients with unexplained pediatric epilepsy and nondiagnostic exome sequencing (ES), genome sequencing (GS) identified five diagnostic and four likely diagnostic genetic findings. Seven of these nine genetic findings required GS for variant detection and could not be detected by current ES methods. CNV: copy number variant, indel: insertion-deletion, SNV: single nucleotide variant.

**Table 2:** Summary of Diagnoses.

| **ID/Sex/Age at seizure onset (years)/Age at genetic diagnosis (years)** | **Clinical features** | **Gene or Cytoband/**<br>**Variant/Zygosity/**<br>**Inheritance/Classification** | **Previous genetic testing** | **Ability to detect in ES vs GS** | **Documented utility** |
| --- | --- | --- | --- | --- | --- |
| 1/F/0-5/11-15 | Combined epilepsy, DD, low average cognitive abilities, ASD, behavioral dysregulation, headaches | CDK19/NM_015076.5:c.386 A>C, p.(K129T)/Heterozygous/Paternal/VUS | CMA, epilepsy panel, ES | Detectable in ES, recognized as likely mosaic in father during analysis of GS |  |
| 2/M/0-5/16-20 | DEE (LGS), GDD, moderate to severe ID, ASD, aggressive behavior, Type 1 diabetes mellitus | SYNGAP1/NM_006772.2:c.2590_2591ins(71), p.(A864Vfs*18)/Heterozygous/De novo, P | Epilepsy panel, ES, mitochondrial sequencing | Filtered out as a sequencing error during analysis of ES due to being in a noisy region | Research study, family support group |
| 3/M/0-5/16-20 | DEE (IESS), GDD, severe ID, ASD, sleep disturbances, behavioral dysregulation, distinctive facial features, contractures, history of malrotation | 11p11.2/chr11:g.46114746_46122325del/Heterozygous/De novo/VUS | CMA, Fragile X, 15q11q13 methylation, epilepsy and ASD panels, ES | Not covered by ES due to being in a noncoding region (overlaps promoter of *PHF21A*) |  |
| 4/F/16-20/21-25 | Combined epilepsy, regression, mild ID, ASD | 12q13.2/chr12:g.56133540_56169966del/Heterozygous/De novo/P | CMA, ES | Detectable in ES with current CNV analysis methods although harder to visualize compared to in GS (includes part of *SMARCC2*) | Research study, family support group |
| 5/F/0-5/6-10 | Combined epilepsy, low average cognitive abilities, specific learning disorder with reading impairment, generalized anxiety disorder | SCN1A/NM_001165963.1:c.4002+2454G>A/Heterozygous/Paternal/VUS | CMA, ES | Not covered by ES due to being in a noncoding region (deep intronic variant in poison exon) | Impact on treatment (ASM choices), foundation |
| 6/F/0-5/0-5 | NAFE | *PRRT2*/NM_145239.2:c.649 dup, p.R217Pfs*8/<br>Heterozygous/Maternal/P | ES | Not called during analysis of ES due to low coverage and being in a difficult to sequence region | Impact on treatment (weaned off ASM given natural history), impact on prognosis (likelihood of normal development, risk of other manifestations (PKD, hemiplegic migraines, episodic ataxia)) |
| 7/F/0-5/0-5 | DEE (IESS), DD, regression, microcephaly, CVI, spastic quadriparesis | *IRF2BPL*/NM_024496.4:c.283_320del, p.(A95Tfs*25)/<br>Heterozygous/De novo/LP | Epilepsy panel, ES | Not called during analysis of ES due to low coverage and being in a repetitive region | Impact on prognosis (e.g., risk of movement disorder), research study, foundation |
| 8/F/0-5/6-10 | DEE, GDD, microcephaly, hypotonia, strabismus, short stature, simplified cortical gyral pattern with ventriculomegaly and thinned corpus callosum, prematurity (born at 34 weeks gestational age) | *RNU4-2*/chr12:g.120291839_120291840insA, n.64_65insT/<br>Heterozygous/De novo/VUS | CMA, epilepsy and brain malformation panels, ES | Not covered by ES due to being in a noncoding gene/exon |  |
| 9/F/0-5/6-10 | NAFE, GDD, hypotonia | *MECP2* exon 4/chrX:g.154030723_154031144del/<br>Heterozygous/De novo/P | CMA, epilepsy panel, ES | Not called during analysis of ES due to small size | Impact on treatment (FDA-approved trofinetide), impact on workup (referral to Rett syndrome clinic, referral to Ophthalmology to evaluate for CVI, EKG, fasting lipid profile), family support group |

ASM: anti-seizure medication, CVI: cortical visual impairment, F: female, LGS: Lennox-Gastaut Syndrome, M: male

Of the five individuals with diagnostic genetic findings, three have P/LP coding SNVs/indels requiring GS for detection, one has a small pathogenic CNV requiring GS for detection, and one has a pathogenic CNV detectable by current ES methods. Patient 2, a boy with Lennox-Gastaut Syndrome, ASD, severe ID, and behavioral dysregulation, has a pathogenic *de novo* indel in *SYNGAP1* (OMIM 603384) that was filtered out during ES analysis due to being in a noisy region and required GS for detection. Patient 4, a woman with combined epilepsy, ASD, and mild ID, has a pathogenic *de novo* deletion of chr12q13.2, including part of *SMARCC2*, missed on previous CMA and ES analysis but detectable using current ES CNV pipelines. Patient 6, a girl with NAFE, has a pathogenic maternally inherited frameshift variant in *PRRT2* (OMIM 614386) that was not called on previous ES analysis due to low coverage and being in a difficult to sequence region and required GS for detection. Patient 7, a girl with Infantile Epileptic Spasms Syndrome (IESS), microcephaly, cortical visual impairment, and spastic quadriparesis, has a *de novo* likely pathogenic indel in *IRF2BPL* (OMIM 611720) that was not called on previous ES analysis due to low coverage and being in a difficult to sequence region and required GS for detection (**Figure 3A**). Patient 9, a girl with NAFE, global DD, and hypotonia, has a *de novo* pathogenic deletion of exon 4 of *MECP2* (OMIM 300005) that was not called on previous CMA or ES analysis due to small size and required GS for detection (**Figure 3B**).

**Figure 3:**
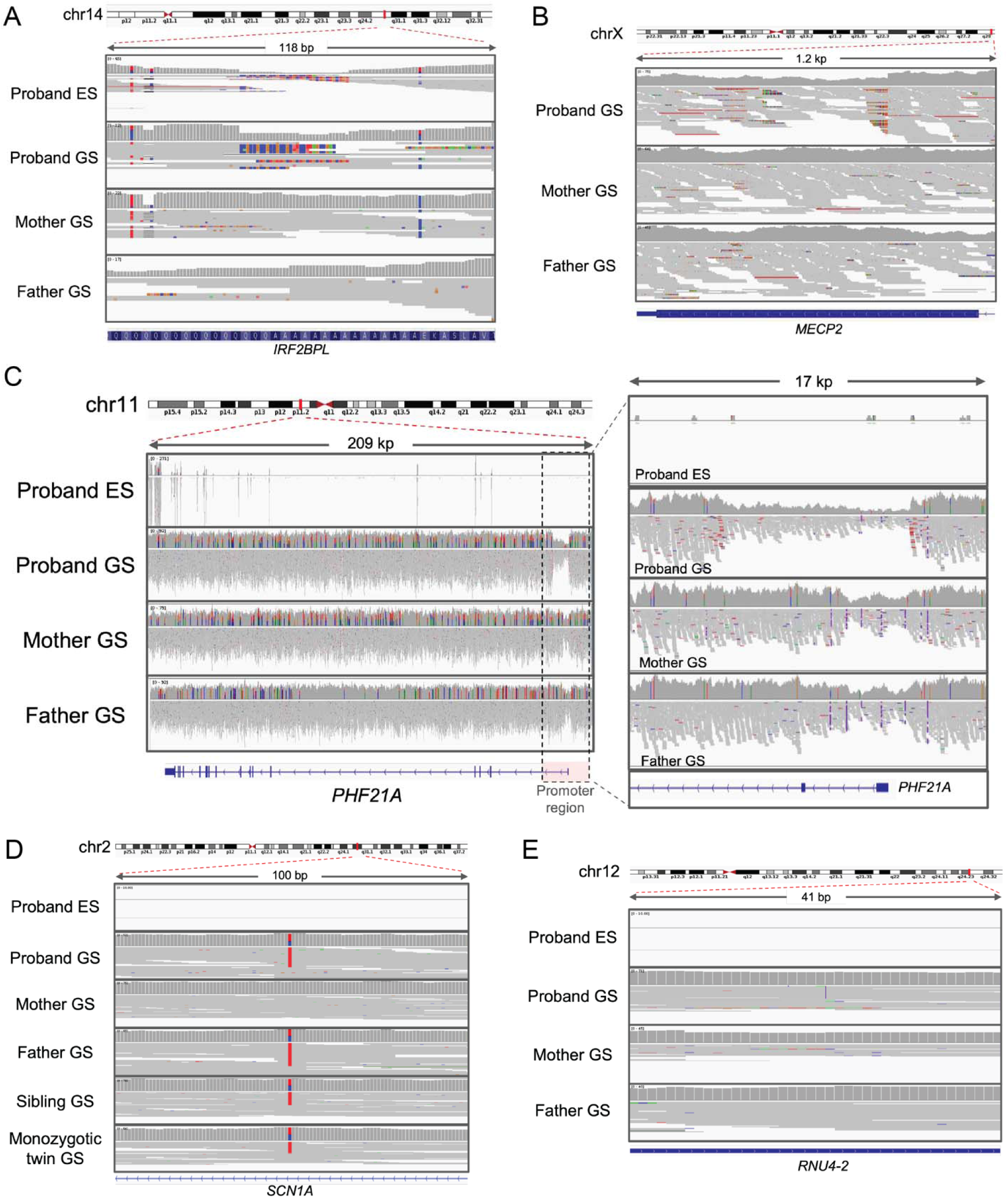
Pathogenic Variants Uniquely Detectable by GS. A: The *de novo* frameshift variant in *IRF2BPL* in Patient 7 is difficult to detect in the ES data due to low coverage and a difficult to sequence region, but it is detectable in the GS data. B: The *de novo* deletion in *MECP2* in Patient 9 is difficult to detect using current ES calling methods due to its small size (for this case, clinical ES data was not available in the research setting), but is detectable in the GS data. C: The *de novo* deletion overlapping the promoter of *PHF21A* in Patient 3 is not covered in the ES data, but is detectable in the GS data. D: The deep intronic variant in an *SCN1A* poison exon in Patient 5, which segregated in her family, is not covered in the ES data, but is detectable in the GS data E: The *de novo* insertion in the noncoding gene *RNU4-2* in Patient 8 is not covered in the ES data, but is detectable in the GS data.

Of the four individuals with likely diagnostic genetic findings, three have noncoding variants that required GS for detection and one has a coding SNV detectable by current ES methods. Patient 1, a girl with combined epilepsy, ASD, DD, low average cognitive abilities, and behavioral dysregulation, has a paternally inherited missense VUS in *CDK19* (OMIM 614720). The paternal variant allele fraction (0.27) is suggestive of mosaicism, and her clinical features fit the *CDK19*-associated epilepsy phenotype. The variant is detectable by ES, and the father’s skewed VAF was recognized during GS analysis. Patient 3, a man with a DEE (IESS), ASD, severe ID, behavioral dysregulation, and dysmorphic features, has a *de novo* deletion of uncertain significance of chr11p11.2 that overlaps the promoter of *PHF21A* (OMIM 608325) and required GS for detection (**Figure 3C**). He has had an extensive nondiagnostic genetic workup through clinical and research efforts, and his clinical features fit the phenotype of *PHF21A*-associated Intellectual Developmental Disorder with Behavioral Abnormalities and Craniofacial Dysmorphism with or without Seizures (OMIM 618725). Patient 5, a girl with Generalized Epilepsy with Febrile Seizures Plus, low average cognitive abilities, learning disorder with reading impairment, and generalized anxiety disorder, has a paternally inherited deep intronic VUS in *SCN1A* (OMIM 182389) that required GS for detection (**Figure 3D**). The variant segregates with her two similarly affected siblings and is in a poison exon near variants previously reported in participants with SCN1A-associated phenotypes, and her affected family members similarly fit the phenotype of SCN1A-associated epilepsies.^36^ Patient 8, a girl with DEE, GDD, microcephaly, small stature, hypotonia, and MRI abnormalities, has a *de novo* insertion of uncertain significance in a noncoding exon of the noncoding RNA *RNU4-2* that is not captured by clinical ES testing and required GS for detection (**Figure 3E**). This insertion was recently reported associated with syndromic neurodevelopmental disorders that fit her phenotype.^37^

### Clinical Utility of GS Diagnoses

The median length of the diagnostic odyssey was 6 years (range 3-16.75) for the nine participants with diagnostic/likely diagnostic genetic findings. In four cases, the genetic findings had documented clinical utility. For Patient 6 (*PRRT2*), the genetic finding led the treating clinician to wean off anti-seizure medication given the natural history of *PRRT2*-associated epilepsy and provided prognostic information regarding the high likelihood of typical development and cognitive abilities as well as the risk of other *PRRT2*-associated manifestations (paroxysmal kinesigenic dyskinesia, hemiplegic migraines, episodic ataxia) to monitor for. For Patient 7 (*IRF2BPL*), the genetic finding provided prognostic information about the risk of movement disorder and contributed to recognition of likely dystonic episodes. For Patient 9 (*MECP2*), the genetic finding led the treating clinician to seek insurance approval for trofinetide and to additional subspecialty referrals (Rett Syndrome clinic, Ophthalmology) and workup (EKG, fasting lipid profile). For Patient 5 (*SCN1A*), the genetic finding influenced treatment and suggested potential precision management approaches.

For three participants, the genetic findings allowed the families to access etiology-specific research studies (*IRF2BPL*, chr12 deletion including *SMARCC2*, and *SYNGAP1*). Further, for five participants, the genetic findings allowed the families to access etiology-specific foundations and/or support groups (*IRF2BPL*, *MECP2, SCN1A*, chr12 deletion including *SMARCC2*, and *SYNGAP1*).

## Discussion

We report the utility of GS after nondiagnostic ES in a rigorously phenotyped cohort of participants with unexplained pediatric epilepsy, including DEE and non-DEE phenotypes. We find that GS can identify variants in coding regions that are missed by current ES methods and variants in noncoding regions that are difficult or not possible to detect by ES. Our comprehensive GS analyses identified five cases with diagnostic genetic findings and four cases with likely diagnostic genetic findings, for a diagnostic yield of GS after ES of 7.2%. For 7/9 cases (78%), GS was required for variant detection, for a unique diagnostic yield of GS of 5.6%. For 4/9 cases (44%), the genetic findings had immediate clinical utility, and for 5/9 cases (56%), the genetic findings had potential personal utility for families through access to etiology-specific foundations/support groups. All nine cases experienced long diagnostic odysseys— median six years—highlighting the importance of evaluating and implementing genomic tests from the research laboratory into clinical epilepsy care. Short-read GS is currently clinically accredited but access in clinical epilepsy care is limited; ideally, rapid GS at the time of epilepsy diagnosis will provide early answers to families and access to emerging precision therapies to optimize outcomes.^35^

Two previous studies of GS after nondiagnostic ES in epilepsy reported smaller cohorts with DEE/EE and performed limited analyses. Palmer and colleagues analyzed GS data from 15 participants with DEE and previous nondiagnostic ES for coding and regulatory SNVs and indels, CNVs, SVs, mitochondrial, and noncoding and deep intronic variants (their methods do not specify the prediction tools and thresholds used to prioritize regulatory, noncoding, and deep intronic variants).^21^ They reported a GS after ES yield of 53% (8/15) and a unique to GS yield of 20% (3/15), specifically three complex structural variants.^21^ Grether and colleagues analyzed GS data from 20 participants with DEE/EE and previous nondiagnostic ES for SNVs, indels, CNVs, and *de novo* variants in the *MEF2C* 5’ UTR.^20^ They reported a GS after ES yield of 20% (4/20) and a unique to GS yield of 0%.^20^ We performed more comprehensive GS analyses in a larger and more phenotypically diverse cohort and report a GS after ES yield of 7.2%, with a unique to GS yield of 5.6%. Our cohort included 44 participants with DEE/EE, for which we report a GS after ES yield of 9.1% (4/44) and a unique to GS yield of 9.1% (4/44). The larger overall yields and variable unique to GS yields in the previous studies are likely due to their smaller cohort sizes. Our findings are consistent with those recently reported in a large heterogenous rare disease cohort, which reported 8.2% (61/744) of their genetic diagnoses required GS (not all participants had previous ES).^38^ We note that our *additional* yield of GS in epilepsy is comparable to the *total* yield of CMA in epilepsy.^14^ We expect the yield of GS to continue to increase as sequencing costs continue to decline and we continue to learn more about the noncoding regions of the human genome in relation to human disease.

### Limitations

Our study has some limitations, including recruiting from a single pediatric referral center, including largely White and non-Hispanic participants (reflecting the demographics of the BCH CRDC^39^ but not the worldwide population of individuals with epilepsy), and including participants referred for research genomic sequencing (patients who qualified for clinical testing may have higher yield). We included participants who did not have both biological parents available, which we felt was important for equitable access to genomic sequencing and mirrors common clinical scenarios, but potentially limited GS interpretation. Further, we abstracted utility from the EMR and had limited ability to assess the impact of genetic diagnoses on participants and their families.^40,41^ Future studies of GS in larger and more diverse cohorts of participants with epilepsy, studies evaluating emerging long-read GS and multi-omics approaches, and studies of prospectively ascertained cohorts assessing a broad definition of utility will provide further insight into the etiologies underlying epilepsy and the impact of identifying those etiologies.

### Conclusions

Our cohort study demonstrates the utility of GS after nondiagnostic ES in participants with unexplained pediatric epilepsy, including participants with DEE and non-DEE phenotypes. Our findings demonstrate that GS can identify genetic diagnoses not detected by ES and support the implementation of GS as a first-line genetic test in clinical epilepsy care in the future.

## Supporting information

Supplementary Methods and References

## Data Availability

Deidentified participant data will be made available on request to the corresponding author. Data will be made available for research purposes to investigators whose protocols allow use of deidentified phenotypic and genomic data with a signed data use agreement.

## Author Contributions

Drs. D’Gama and Poduri had full access to all the data in the study and take responsibility for the integrity of the data and the accuracy of the data analysis.

*Study concept and design:* D’Gama, Shao, Rockowitz, Poduri

*Acquisition, analysis, or interpretation of data:* All authors

*Drafting of the manuscript:* D’Gama, Shao

*Critical revision of the manuscript for important intellectual content:* All authors

*Statistical analysis:* D’Gama, Shao

*Obtained funding:* D’Gama, Poduri

*Administrative, technical, or material support:* All authors

*Study supervision:* Poduri

## Additional Contributions

We thank the participants and their families who participated in this study. We thank the BCH clinicians who referred participants to this study, including Christelle Moufawad El Achkar, MD (Department of Neurology, BCH and HMS), Chellamani Harini, MD (Department of Neurology, BCH and HMS), Phillip L. Pearl, MD (Division of Epilepsy and Clinical Neurophysiology, BCH; Department of Neurology, HMS) and Jurriaan M. Peters, MD, PhD (Department of Neurology, BCH and HMS). We thank Courtney E. French, PhD (CRDC, BCH) and Piotr Sliz, PhD (CRDC and Division of Molecular Medicine, BCH; Departments of Pediatrics and Biological Chemistry and Molecular Pharmacology, HMS) for assistance with data management and review of the manuscript. None of the contributors were compensated for their work.

## References

1. Fisher RS, Acevedo C, Arzimanoglou A, et al. ILAE official report: a practical clinical definition of epilepsy. Epilepsia. Apr 2014;55(4):475–82. doi:10.1111/epi.12550

2. Wei SH, Lee WT. Comorbidity of childhood epilepsy. J Formos Med Assoc. Nov 2015;114(11):1031–8. doi:10.1016/j.jfma.2015.07.015

3. Hesdorffer DC, Logroscino G, Benn EK, Katri N, Cascino G, Hauser WA. Estimating risk for developing epilepsy: a population-based study in Rochester, Minnesota. Neurology. Jan 4 2011;76(1):23–7. doi:10.1212/WNL.0b013e318204a36a

4. Auvin S, Galanopoulou AS, Moshe SL, et al. Revisiting the concept of drug-resistant epilepsy: A TASK1 report of the ILAE/AES Joint Translational Task Force. Epilepsia. Nov 2023;64(11):2891–2908. doi:10.1111/epi.17751

5. Specchio N, Wirrell EC, Scheffer IE, et al. International League Against Epilepsy classification and definition of epilepsy syndromes with onset in childhood: Position paper by the ILAE Task Force on Nosology and Definitions. Epilepsia. Jun 2022;63(6):1398–1442. doi:10.1111/epi.17241

6. Zuberi SM, Wirrell E, Yozawitz E, et al. ILAE classification and definition of epilepsy syndromes with onset in neonates and infants: Position statement by the ILAE Task Force on Nosology and Definitions. Epilepsia. Jun 2022;63(6):1349–1397. doi:10.1111/epi.17239

7. Berg AT, Coryell J, Saneto RP, et al. Early-Life Epilepsies and the Emerging Role of Genetic Testing. JAMA Pediatr. Sep 1 2017;171(9):863–871. doi:10.1001/jamapediatrics.2017.1743

8. Howell KB, Freeman JL, Mackay MT, et al. The severe epilepsy syndromes of infancy: A population-based study. Epilepsia. Feb 2021;62(2):358–370. doi:10.1111/epi.16810

9. Symonds JD, Elliott KS, Shetty J, et al. Early childhood epilepsies: epidemiology, classification, aetiology, and socio-economic determinants. Brain. Oct 22 2021;144(9):2879–2891. doi:10.1093/brain/awab162

10. McTague A, Howell KB, Cross JH, Kurian MA, Scheffer IE. The genetic landscape of the epileptic encephalopathies of infancy and childhood. Lancet Neurol. Mar 2016;15(3):304–16. doi:10.1016/S1474-4422(15)00250-1

11. Symonds JD, McTague A. Epilepsy and developmental disorders: Next generation sequencing in the clinic. Eur J Paediatr Neurol. Jan 2020;24:15–23. doi:10.1016/j.ejpn.2019.12.008

12. Kim J, Hu C, Moufawad El Achkar C, et al. Patient-Customized Oligonucleotide Therapy for a Rare Genetic Disease. N Engl J Med. Oct 24 2019;381(17):1644–1652. doi:10.1056/NEJMoa1813279

13. Koh HY, Smith L, Wiltrout KN, et al. Utility of Exome Sequencing for Diagnosis in Unexplained Pediatric-Onset Epilepsy. JAMA Netw Open. Jul 3 2023;6(7):e2324380. doi:10.1001/jamanetworkopen.2023.24380

14. Sheidley BR, Malinowski J, Bergner AL, et al. Genetic testing for the epilepsies: A systematic review. Epilepsia. Feb 2022;63(2):375–387. doi:10.1111/epi.17141

15. Epi KC, Epilepsy Phenome/Genome P, Allen AS, et al. De novo mutations in epileptic encephalopathies. Nature. Sep 12 2013;501(7466):217-21. doi:10.1038/nature12439

16. Epi25 Collaborative. Electronic address sbuea, Epi C. Ultra-Rare Genetic Variation in the Epilepsies: A Whole-Exome Sequencing Study of 17,606 Individuals. Am J Hum Genet. Aug 1 2019;105(2):267–282. doi:10.1016/j.ajhg.2019.05.020

17. Oliver KL, Scheffer IE, Bennett MF, Grinton BE, Bahlo M, Berkovic SF. Genes4Epilepsy: An epilepsy gene resource. Epilepsia. May 2023;64(5):1368–1375. doi:10.1111/epi.17547

18. Wojcik MH, Reuter CM, Marwaha S, et al. Beyond the exome: What’s next in diagnostic testing for Mendelian conditions. Am J Hum Genet. Aug 3 2023;110(8):1229–1248. doi:10.1016/j.ajhg.2023.06.009

19. Smith L, Malinowski J, Ceulemans S, et al. Genetic testing and counseling for the unexplained epilepsies: An evidence-based practice guideline of the National Society of Genetic Counselors. J Genet Couns. Apr 2023;32(2):266–280. doi:10.1002/jgc4.1646

20. Grether A, Ivanovski I, Russo M, et al. The current benefit of genome sequencing compared to exome sequencing in patients with developmental or epileptic encephalopathies. Mol Genet Genomic Med. May 2023;11(5):e2148. doi:10.1002/mgg3.2148

21. Palmer EE, Sachdev R, Macintosh R, et al. Diagnostic Yield of Whole Genome Sequencing After Nondiagnostic Exome Sequencing or Gene Panel in Developmental and Epileptic Encephalopathies. Neurology. Mar 30 2021;96(13):e1770–e1782. doi:10.1212/WNL.0000000000011655

22. Harris PA, Taylor R, Thielke R, Payne J, Gonzalez N, Conde JG. Research electronic data capture (REDCap)--a metadata-driven methodology and workflow process for providing translational research informatics support. J Biomed Inform. Apr 2009;42(2):377–81. doi:10.1016/j.jbi.2008.08.010

23. Hirsch E, French J, Scheffer IE, et al. ILAE definition of the Idiopathic Generalized Epilepsy Syndromes: Position statement by the ILAE Task Force on Nosology and Definitions. Epilepsia. Jun 2022;63(6):1475–1499. doi:10.1111/epi.17236

24. Riney K, Bogacz A, Somerville E, et al. International League Against Epilepsy classification and definition of epilepsy syndromes with onset at a variable age: position statement by the ILAE Task Force on Nosology and Definitions. Epilepsia. Jun 2022;63(6):1443–1474. doi:10.1111/epi.17240

25. Scheffer IE, Berkovic S, Capovilla G, et al. ILAE classification of the epilepsies: Position paper of the ILAE Commission for Classification and Terminology. Epilepsia. Apr 2017;58(4):512–521. doi:10.1111/epi.13709

26. Rockowitz S, LeCompte N, Carmack M, et al. Children’s rare disease cohorts: an integrative research and clinical genomics initiative. NPJ Genom Med. 2020;5:29. doi:10.1038/s41525-020-0137-0

27. Dolzhenko E, van Vugt J, Shaw RJ, et al. Detection of long repeat expansions from PCR-free whole-genome sequence data. Genome Res. Nov 2017;27(11):1895–1903. doi:10.1101/gr.225672.117

28. Laricchia KM, Lake NJ, Watts NA, et al. Mitochondrial DNA variation across 56,434 individuals in gnomAD. Genome Res. Mar 2022;32(3):569–582. doi:10.1101/gr.276013.121

29. Lee E, Iskow R, Yang L, et al. Landscape of somatic retrotransposition in human cancers. Science. Aug 24 2012;337(6097):967–71. doi:10.1126/science.1222077

30. McKenna A, Hanna M, Banks E, et al. The Genome Analysis Toolkit: a MapReduce framework for analyzing next-generation DNA sequencing data. Genome Res. Sep 2010;20(9):1297–303. doi:10.1101/gr.107524.110

31. Felker SA, Lawlor JMJ, Hiatt SM, et al. Poison exon annotations improve the yield of clinically relevant variants in genomic diagnostic testing. Genet Med. Aug 2023;25(8):100884. doi:10.1016/j.gim.2023.100884

32. Karczewski KJ, Francioli LC, Tiao G, et al. The mutational constraint spectrum quantified from variation in 141,456 humans. Nature. May 2020;581(7809):434–443. doi:10.1038/s41586-020-2308-7

33. Richards S, Aziz N, Bale S, et al. Standards and guidelines for the interpretation of sequence variants: a joint consensus recommendation of the American College of Medical Genetics and Genomics and the Association for Molecular Pathology. Genet Med. May 2015;17(5):405–24. doi:10.1038/gim.2015.30

34. Riggs ER, Andersen EF, Cherry AM, et al. Technical standards for the interpretation and reporting of constitutional copy-number variants: a joint consensus recommendation of the American College of Medical Genetics and Genomics (ACMG) and the Clinical Genome Resource (ClinGen). Genet Med. Feb 2020;22(2):245–257. doi:10.1038/s41436-019-0686-8

35. D’Gama AM, Mulhern S, Sheidley BR, et al. Evaluation of the feasibility, diagnostic yield, and clinical utility of rapid genome sequencing in infantile epilepsy (Gene-STEPS): an international, multicentre, pilot cohort study. Lancet Neurol. Sep 2023;22(9):812–825. doi:10.1016/S1474-4422(23)00246-6

36. Carvill GL, Engel KL, Ramamurthy A, et al. Aberrant Inclusion of a Poison Exon Causes Dravet Syndrome and Related SCN1A-Associated Genetic Epilepsies. Am J Hum Genet. Dec 6 2018;103(6):1022–1029. doi:10.1016/j.ajhg.2018.10.023

37. Chen Y, Dawes R, Kim HC, et al. De novo variants in the non-coding spliceosomal snRNA gene RNU4-2 are a frequent cause of syndromic neurodevelopmental disorders. medRxiv. 2024;doi:10.1101/2024.04.07.24305438

38. Wojcik MH, Lemire G, Zaki MS, et al. Unique Capabilities of Genome Sequencing for Rare Disease Diagnosis. medRxiv. Aug 13 2023;doi:10.1101/2023.08.08.23293829

39. Frazier ZJ, Brown E, Rockowitz S, et al. Toward representative genomic research: the children’s rare disease cohorts experience. Ther Adv Rare Dis. Jan-Dec 2023;4:26330040231181406. doi:10.1177/26330040231181406

40. Hayeems RZ, Luca S, Assamad D, Bhatt A, Ungar WJ. Utility of Genetic Testing from the Perspective of Parents/Caregivers: A Scoping Review. Children (Basel). Mar 27 2021;8(4)doi:10.3390/children8040259

41. Smith HS, Morain SR, Robinson JO, et al. Perceived Utility of Genomic Sequencing: Qualitative Analysis and Synthesis of a Conceptual Model to Inform Patient-Centered Instrument Development. Patient. May 2022;15(3):317–328. doi:10.1007/s40271-021-00558-4

