## Supplementary Methods and References for "Utility of Genome Sequencing After Nondiagnostic Exome Sequencing in Unexplained Pediatric Epilepsy"

**Table of Contents**

eMethods 2

eReferences 5

### eMethods

#### Genome Sequencing

At the CLIA-certified laboratory (GeneDx),[1] DNA was extracted from blood samples using standard methods and sequencing libraries were prepared using Illumina DNA PCR-Free Prep, Tagmentation. Paired end (2 x 150 bp) GS was performed on Illumina NovaSeq 6000 platforms with average depth of 43X. Raw sequencing data in fastq format were securely transferred to BCH for analyses.

#### Variant Calling, Annotation, and Filtering

Raw sequencing data were aligned to the hg38 genome using DRAGEN v3.9.3.

Coding Single Nucleotide Variants and Small Insertions-Deletions

Coding SNVs and indels were identified using DRAGEN v3.9.3 small variant caller and analyzed via GeneDx Discovery Platform and Illumina Emedgene Analyze. Rare, potentially clinically relevant variants were identified based on the following criteria:

a) variant overlaps coding region of an epilepsy-associated gene (based on OMIM and PubMed search)
b) gnomAD AF <= 0.1%[2]
c) not ClinVar B/LB[3]
d) Zygosity and inheritance consistent with human disease association (i.e., for a disease with autosomal dominant mode of inheritance, heterozygous variant is de novo or pseudo de novo if one or both biological parents data available for analysis respectively or heterozygous variant is inherited and segregates with affected family members if family members data available or heterozygous variant impacts a gene with sufficient evidence of incomplete penetrance; for a disease with autosomal recessive mode of inheritance, variant is homozygous or hemizygous or variants are compound heterozygous)

Copy Number Variants and Structural Variants:

CNVs and SVs were identified using DRAGEN v3.9.3 CNV and SV caller. Resulting CNVs and SVs were processed and annotated via an in-house bioinformatics pipeline. Variant quality, inheritance pattern, impact on gene, overlap to known pathogenic variant in ClinVar, overlap to DECIPHER recurrent CNV syndromes,[4] frequency information from gnomAD SV database, and frequency information calculated using BCH Children's Rare Disease Cohorts (CRDC) data[5] were utilized for variant prioritization. Rare, potentially clinically relevant variants were identified based on the following criteria:

a) variant overlaps recurrent CNV syndrome region or exon of an epilepsy-associated gene or is a large CNV >1 MB
b) CRDC AF <= 0.5% for dominant inheritance, CRDC AF <= 1% for recessive inheritance, and gnomAD AF <= 0.5%
c) CRDC homozygous count (for autosomal recessive) or hemizygous count (for X-linked) <= 8
d) not ClinVar B/LB
e) Zygosity and inheritance consistent with human disease association as above

Short Tandem Repeats:

Short tandem repeat expansions were identified using ExpansionHunter v5.0.0.[6] The variant catalog used in short tandem repeat quantification included all known repeat disease loci curated by Illumina (https://github.com/Illumina/ExpansionHunter/blob/master/variant_catalog/hg38/variant_catalog.json) and the location of polyalanine tract 1 and 2 at ARX. Repeat expansion results were processed and annotated via an in-house bioinformatics pipeline. Inheritance pattern, known pathological expansion threshold, frequency information from gnomAD short tandem repeat database, and frequency information from all GS datasets in this epilepsy cohort were utilized for variant prioritization. Rare, potentially clinically relevant repeat expansions were identified based on the following criteria:

a) repeat number passes known pathologic threshold
b) internal cohort AF <= 5% and gnomAD AF <= 5%
c) zygosity and inheritance consistent with human disease association as above

Mobile Element Insertions:

Mobile Element insertions were identified using xTEA v0.1.9.[7] The resulting Alu, HERV, LINE1, and SVA insertions were merged for downstream analysis. An in-house bioinformatics pipeline was utilized to annotate mobile element insertions with inheritance pattern, impact on gene, and frequency information from gnomAD SV database, 1000G SV database,[8] and all GS datasets in this epilepsy cohort. Rare, potentially clinically relevant variants were identified based on the following criteria:

a) variant overlaps exon of an epilepsy-associated gene
b) internal cohort AF <= 5%, gnomAD AF <= 1%, and 1000G AF <= 1%
c) internal cohort homozygous count (for autosomal recessive) or hemizygous count (for X-linked) <= 4
d) zygosity and inheritance consistent with human disease association as above

Mitochondrial Variants:

Mitochondrial variants were called using a Mutect2-based pipelined adapted from the gnomAD mitochondria workflow.[9] VEP v106 was utilized to annotate mitochondrial variants with impact on gene, PolyPhen Score, and SIFT score.[10] An in-house bioinformatics pipeline was then applied to add MITOMAP disease and polymorphism, inheritance pattern, frequency calculation from gnomAD database, and frequency calculation from BCH CRDC data. Rare, potentially clinically relevant variants were identified based on the following criteria:

a) variant overlaps an epilepsy-associated gene
b) alternative allele read depth >= 5 and alternative allele VAF >= 1%
c) CRDC AF <= 1% and gnomAD AF <= 1%
d) inheritance is de novo, inherited from affected mother, or no maternal data was available

Noncoding SNVs and indels:

Noncoding SNVs and indels called by DRAGEN small variant caller were filtered based on their genomic location. Variants within 5 kb of exons of GENCODE protein-coding genes or overlapping ENCODE[11] and ENSEMBL[10] cis-regulatory regions were retained for downstream analysis. These variants were further filtered based on variant calling quality and sequencing read quality around variant location. VEP v106 was utilized to add CADD score, GERP score, spliceAI score, gene impact, gnomAD frequency, and ClinVar occurrence. Rare, potentially clinically relevant variants were identified based on the following criteria:

a) variant within 5kb of an epilepsy-associated gene
b) read depth >= 10 and VAF for heterozygous variants between 0.2 to 0.8
c) GERP score >= 10 and CADD PHRED score >= 10
d) spliceAI score >= 0.8 or ClinVar P/LP
e) gnomAD AF <= 0.1%
f) not ClinVar B/LB
g) zygosity and inheritance consistent with human disease association as above

In addition, we retained variants in poison exon cassettes and further filtered as previously described.[12] We also filtered for variants in an 18bp region of *RNU4-2* as previously described.[13]

For each candidate variant, the sequencing reads were visually reviewed using the Integrative Genome Visualizer (IGV) to assess for sequencing artifacts.[14] Short tandem repeat candidates were additionally inspected using REViewer (https://github.com/Illumina/REViewer).

**References**


1. Retterer K, Juusola J, Cho MT, et al. Clinical application of whole-exome sequencing across clinical indications. *Genet Med*. Jul 2016;18(7):696-704. doi:10.1038/gim.2015.148
2. Karczewski KJ, Francioli LC, Tiao G, et al. The mutational constraint spectrum quantified from variation in 141,456 humans. *Nature*. May 2020;581(7809):434-443. doi:10.1038/s41586-020-2308-7
3. Landrum MJ, Lee JM, Riley GR, et al. ClinVar: public archive of relationships among sequence variation and human phenotype. *Nucleic Acids Res*. Jan 2014;42(Database issue):D980-5. doi:10.1093/nar/gkt1113
4. Firth HV, Richards SM, Bevan AP, et al. DECIPHER: Database of Chromosomal Imbalance and Phenotype in Humans Using Ensembl Resources. *Am J Hum Genet*. Apr 2009;84(4):524-33. doi:10.1016/j.ajhg.2009.03.010
5. Rockowitz S, LeCompte N, Carmack M, et al. Children's rare disease cohorts: an integrative research and clinical genomics initiative. *NPJ Genom Med*. 2020;5:29. doi:10.1038/s41525-020-0137-0
6. Dolzhenko E, van Vugt J, Shaw RJ, et al. Detection of long repeat expansions from PCR-free whole-genome sequence data. *Genome Res*. Nov 2017;27(11):1895-1903. doi:10.1101/gr.225672.117
7. Lee E, Iskow R, Yang L, et al. Landscape of somatic retrotransposition in human cancers. *Science*. Aug 24 2012;337(6097):967-71. doi:10.1126/science.1222077
8. Genomes Project C, Auton A, Brooks LD, et al. A global reference for human genetic variation. *Nature*. Oct 1 2015;526(7571):68-74. doi:10.1038/nature15393
9. Laricchia KM, Lake NJ, Watts NA, et al. Mitochondrial DNA variation across 56,434 individuals in gnomAD. *Genome Res*. Mar 2022;32(3):569-582. doi:10.1101/gr.276013.121
10. Martin FJ, Amode MR, Aneja A, et al. Ensembl 2023. *Nucleic Acids Res*. Jan 6 2023;51(D1):D933-D941. doi:10.1093/nar/gkac958
11. Consortium EP, Moore JE, Purcaro MJ, et al. Expanded encyclopaedias of DNA elements in the human and mouse genomes. *Nature*. Jul 2020;583(7818):699-710. doi:10.1038/s41586-020-2493-4
12. Felker SA, Lawlor JMJ, Hiatt SM, et al. Poison exon annotations improve the yield of clinically relevant variants in genomic diagnostic testing. *Genet Med*. Aug 2023;25(8):100884. doi:10.1016/j.gim.2023.100884
13. Chen Y, Dawes R, Kim HC, et al. De novo variants in the non-coding spliceosomal snRNA gene RNU4-2 are a frequent cause of syndromic neurodevelopmental disorders. *medRxiv*. 2024;doi:10.1101/2024.04.07.24305438
14. Robinson JT, Thorvaldsdottir H, Winckler W, et al. Integrative genomics viewer. *Nat Biotechnol*. Jan 2011;29(1):24-6. doi:10.1038/nbt.1754
